# Common genetic variant of GC associated with vitamin D deficiency in a Chinese population in the Netherlands

**DOI:** 10.1101/2022.08.08.22278561

**Authors:** Ping Wai Man, Stefan Böhringer, Elisa J.F. Houwink, Wenzhi Lin, Mattijs E. Numans, Paul Lips, Barend J. C. Middelkoop

## Abstract

**Background:** Genome-wide association studies among European populations have identified four single nucleotide polymorphisms (SNPs) in genes involved in vitamin D transport and metabolism affecting 25-hydroxyvitamin D [25(OH)D] concentration: rs2282679 in GC, rs6013897 near CYP24A1, rs10741657 near CYP2R1, and rs12785878 near DHCR7. It is, however, unknown whether the association with 25(OH)D can also be observed in a Chinese population living in the Netherlands.

**Methods:** Observational study. Analyses were performed under an additive (univariate and multivariate) and genotypic model using logistic regression.

**Results:** The C allele (minor allele frequency 0.28) of rs2282679 in GC was associated with vitamin D deficiency as defined by the Health Council of the Netherlands: 25(OH)D <30 nmol/l (or <50 nmol/l for persons aged ≥70 years). Univariate and multivariate odds ratios, 1.52 (95% CI 1.03, 2.24) and 1.57 (95% CI 1.04, 2.39), respectively, were not significant after Bonferroni correction. However, after extending the data by a meta-analysis, a significant association between C allele of rs2282679 and vitamin D deficiency (< 50 nmol/l) was observed (odds ratio 1.33; 95% CI 1.16, 1.52).

**Conclusion:** Our findings suggest that, apart from sun exposure, lifestyle, and environmental factors, a common variant of GC may be associated with vitamin D deficiency.

## Introduction

Vitamin D is a fat-soluble vitamin synthetized in the skin when exposed to ultraviolet B radiation from sunlight from its precursor 7-dehydrocholesterol (pro-vitamin D). It undergoes hydroxylation in the liver by the enzyme vitamin D 25-hydroxylase (CYP2R1) into its major circulating, but biologically inactive, metabolite 25-hydroxyvitamin D [25(OH)D]. A further hydroxylation in the kidney by the enzyme 25(OH)D-1α-hydroxylase (CYP27B1) produces its biologically active form 1,25-dihydroxyvitamin D [1,25(OH)_2_D] [1].

Vitamin D deficiency is common worldwide and is associated with adverse health outcomes [2-4]. The prevalence of hypovitaminosis D tends to be higher in Asian than in Caucasian populations [5, 6]. Although skin pigmentation, demographics, season of measurement, geographic latitude, lifestyle and cultural habits across populations partly could explain these differences, twin and family studies have suggested that genetic factors may also contribute to variability in 25(OH)D concentration. Heritability estimates of serum 25(OH)D concentrations varied worldwide from 20 to 80% [7-10]. By contrast, the overall estimate of heritability of serum 25(OH)D concentrations attributable to single-nucleotide polymorphisms (SNPs) as identified in genome-wide association studies (GWASs) and conducted among populations of European ancestry was only 7.5% [11].

SNPs at four loci in or near genes involved in vitamin D transport and metabolism associated with 25(OH)D concentrations were identified in GWASs: group-specific component (GC) (vitamin D-binding protein), the 24-hydroxylase gene (CYP24A1), the 25-hydroxylase gene (CYP2R1) and the 7-dehydrocholesterol reductase gene (DHCR7). Whereas GC encodes vitamin D-binding protein involved in transport of 25(OH)D, CYP24A1 encodes 24-hydroxylase that initiates degradation of 25(OH)D and 1,25(OH)_2_D, CYP2R1 encodes vitamin D 25-hydroxylase involved in hepatic 25-hydroxylation, and DHCR7 encodes the enzyme 7-dehydrocholesterol reductase which converts the vitamin D precursor 7-dehydrocholesterol into cholesterol, thereby removing substrate for synthesis of vitamin D. Strong genome-wide associations with 25(OH)D concentrations were found at three loci in the discovery cohorts, and were confirmed in the replication cohorts: rs2282679 in GC, rs10741657 near CYP2R1, and rs12785878 near DHCR7 [12, 13]. An additional locus, rs6013897 near CYP24A1, was genome-wide significant in the pooled sample [13]. In a recently conducted expanded GWAS, an association between all four loci and 25(OH)D concentration was found [11].

Associations of the genetic variants influencing 25(OH)D levels were all identified in populations of European ancestry. The objective of this study was to explore associations in a sample of 418 Chinese adults living in the Netherlands. As associations with vitamin D deficiency depend on the threshold of 25(OH)D used, and since there is no consensus on the optimal threshold, we explored data using two cut-off points of 25(OH)D for deficiency: according to the recommendations of the Health Council of the Netherlands (HCN), i.e. 30 nmol/l for persons <70 years and 50 nmol/l for persons ≥70 years [14], and 50 nmol/l which most government guidelines recommend as the threshold for bone health [15].

## Materials and Methods

### Study population

This study was part of an observational study on the vitamin D status in a Chinese population in the Netherlands and described in details elsewhere [6]. Briefly, 418 persons aged ≥18 years with a Chinese background (i.e. when they, or at least one parent, were born in mainland China, Hong Kong or Taiwan) and living in the Netherlands participated in the study. Participants were recruited through Chinese welfare, elderly and women organizations and social media in the four cities of the Netherlands with the largest Chinese communities (Rotterdam, Amsterdam, The Hague, and Utrecht).

This study was conducted according to the guidelines laid down in the Declaration of Helsinki and all procedures involving research study participants were approved by the Medical Ethical Committee of the Leiden University Medical Center (approval number P13.279). Written informed consent was obtained from all participants.

### SNP selection and genotyping

We selected four SNPs based on previous GWASs [11-13]. Baseline measurements of blood samples were obtained in March 2014. After overnight fasting, blood was drawn for DNA isolation and genotyping, and measurement of 25(OH)D. DNA was isolated using standard salting-out procedures at the Erasmus MC laboratory Rotterdam (the Human Genotyping Facility, HuGE-F, at the department of Internal Medicine). For genotyping, validated Taqman assays from ThermoFisher were used for all SNPs (rs12785878, rs10741657, rs6013897, and rs2282679). Genotyping reactions were performed in a 384-wells format containing 2 ng DNA, 1x Taqman assay (ThermoFisher) and 1 x genotyping master mix (ABgene). PCR cycling consisted of initial denaturation for 10 min at 95°C, and 40 cycles with denaturation of 15 sec at 95°C and annealing and extension for 60 sec at 60°C. Signals were read with the Taqman 7900HT (Applied Biosystems Inc.) and analysed using the sequence detection system 2.3 software (Applied Biosystems Inc.). To evaluate genotyping accuracy, 16 (4%) random samples were re-genotyped, leading to identical genotypes in all cases.

### Measurement of serum 25(OH)D levels

Serum 25(OH)D concentrations were measured by isotope dilution/online solid-phase extraction liquid chromatography-tandem mass spectrometry (ID-XLC-MS/MS) [16] at the Endocrine Laboratory of the Amsterdam UMC, Vrije Universiteit Amsterdam. The limit of quantitation (LOQ) was 4.0 nmol/l, intra-assay coefficient of variation (CV) was <6%, and inter-assay CV was <8% for concentrations of 25-180 nmol/l.

### Anthropometric measurements

Height (cm) and weight (kg) were measured in standing position and wearing light clothing and no shoes. The body mass index (BMI, kg/m²) was calculated as body weight divided by squared body height. Waist circumference (WC) was measured with a flexible tape all around the body at the level of the navel over thin clothing.

Physical activity was self-reported as a binary variable and defined as a moderate strenuous effort of at least 30 min per day, e.g., walking, cycling, or gardening, or activity below that level.

## Statistical Analysis

### Primary analysis

The characteristics of the participants are presented according to cut-off points of 25(OH)D as recommended by the HCN and by the majority of government guidelines (50 nmol/l). Continuously distributed variables were compared by One-way ANOVA and categorical variables using Chi-square test. Deviation from Hardy-Weinberg equilibrium (HWE) was assessed by a goodness-of-fit Chi-square test. To examine the associations between genetic variants of vitamin D and vitamin D deficiency, also the aforementioned two cut-off points were used.

Analyses of genetic variants of vitamin D were performed under an additive model (using the count of one of the alleles as continuous covariate) as well as a genotypic model (individual effect of each genotype with respect to one homozygous reference genotype). As our main analysis, univariate analysis was performed to separately examine the SNPs (continuous variables) as predictors of vitamin D deficiency. In total, four SNPs were analysed. To evaluate the joint effects of SNPs, we also conducted multivariate analyses including all SNPs (independent variables) in a single model. Initially, genotypic, univariate and multivariate models were adjusted for age, sex, body mass index (BMI) and physical activity. A second model, without BMI and physical activity as covariates, was very similar to the fully adjusted models and, therefore, we only present this simpler model.

### Meta-analysis

For the meta-analysis, a search for suitable studies was performed in electronic databases PubMed and Web of Science up to November 13 2019, using key terms “vitamin D binding protein” and “Chinese” with the combination of “vitamin D deficiency”. As a result, two studies were selected and the odds ratio (OR) of our finding of the C-allele of rs2282679 in GC for the association with vitamin D deficiency [25(OH)D <50 nmol/l] was meta-analysed (fixed-effect model) with those of Chinese people living in Hong Kong [17] and Dali (south-west China) [18].

Results are expressed as OR with their corresponding 95% confidence interval (CI), or *P*-values. As a nominal level, a *P*-value of <0.05 was considered to be statistically significant. To account for multiple testing, a *P*-value of 0.004 (0.05/12) was used as a significance threshold to determine overall significance using Bonferroni correction for testing 12 comparisons. Analyses were performed using IBM SPSS Statistics for Windows, Version 23 (IBM Corp. Arnouk, NY, USA), and the Open Meta-Analyst software, Brown University, Providence, RI, USA [19].

## Results

Our study included a total of 418 individuals with a Chinese background of whom 416 completed serum 25(OH)D measurement (103 men and 313 women); mean age was 56.3 ± 12.1 years. Men had lower median serum 25(OH)D concentration than women (42.0 and 47.0 nmol/l, respectively, *P* = 0.046) and higher BMI (25.4 and 23.9 kg/m^2^, respectively, *P* <0.001) (data not shown). Table 1 shows the characteristics of the participants according to two different cut-off points for vitamin D deficiency: the criteria of the HCN (30 nmol/l for persons <70 years; 50 nmol/l for persons ≥70 years), and a general cut-off of 50 nmol/l (as used in most government guidelines). The prevalence of vitamin D deficiency according to the criteria of the HCN was observed in 23% of the population, and in almost 57% when vitamin D deficiency was defined as 25(OH)D <50 nmol/l. Participants with serum 25(OH)D concentrations ≥50 nmol/l were older and reported significantly more physical activity (≥3 days per week moderate strenuous effort for at least 30 min per day) compared to participants with serum 25(OH)D levels <50 nmol/l (Table 1).

**Table 1.** Characteristics of the participants according to two different cut-off points for vitamin D deficiency

|  | 25(OH)D cut-off point for vitamin D deficiency |  |  |  |  |  |
| --- | --- | --- | --- | --- | --- | --- |
|  | vitamin D deficient | not vitamin D deficient | *P* | vitamin D deficient | not vitamin D deficient | *P* |
|  | 25(OH)D <30 (<70 y)<br>or <50 (≥70 y) nmol/l | 25(OH)D ≥30 (<70 y)<br>or ≥50 (≥ 70 y) nmol/l |  | 25(OH)D <50<br>nmol/l | 25(OH)D ≥50 nmol/l |  |
| Age (years) | 57.0 (14.1) | 56.2 (11.5) | 0.599 | 54.9 (12.9) | 58.4 (10.9) | 0.004 |
| Sex |  |  |  |  |  |  |
| Men (%) | 24 (23.5) | 78 (76.5) | 0.892 | 69 (67.6) | 33 (32.4) | 0.009 |
| Women (%) | 70 (22.9) | 236 (77.1) |  | 162 (52.9) | 144 (47.1) |  |
| Weight (kg) | 61.6 (10.9) | 60.8 (10.5) | 0.517 | 61.7 (10.7) | 60.1 (10.3) | 0.123 |
| Height (cm) | 159.0 (8.0) | 158.2 (7.5) | 0.408 | 159.2 (8.0) | 157.2 (7.0) | 0.008 |
| BMI (kg/m<sup>2</sup>) | 24.3 (3.3) | 24.3 (3.6) | 0.882 | 24.3 (3.2) | 24.3 (3.9) | 0.911 |
| Physical activity |  |  |  |  |  |  |
| < 3 days/week (%) | 54 (22.3) | 188 (77.7) | 0.705 | 152 (62.8) | 90 (37.2) | 0.003 |
| ≥ 3 days/week (%) | 39 (23.9) | 124 (76.1) |  | 78 (47.9) | 85 (52.1) |  |

Data are expressed as numbers (%) or means (SD)
BMI *body mass index*

Invalid measurements of DNA were observed in 46 (11.0%) of the samples of rs2282679, 34 (8.2%) of rs6013897, 37 (8.9%) of rs10741657, and 50 (12.0%) of rs12785878 (data not shown).

The frequencies of rs2282679, rs6013897, and rs10741657 did not deviate from HWE proportions (*P*_*HWE*_ = 0.169, *P*_*HWE*_ = 0.758, *P*_*HWE*_ = 0.974, respectively); however, rs12785878 deviated significantly (*P*_*HWE*_ = 7.120e-5) (Table 2).

**Table 2.**
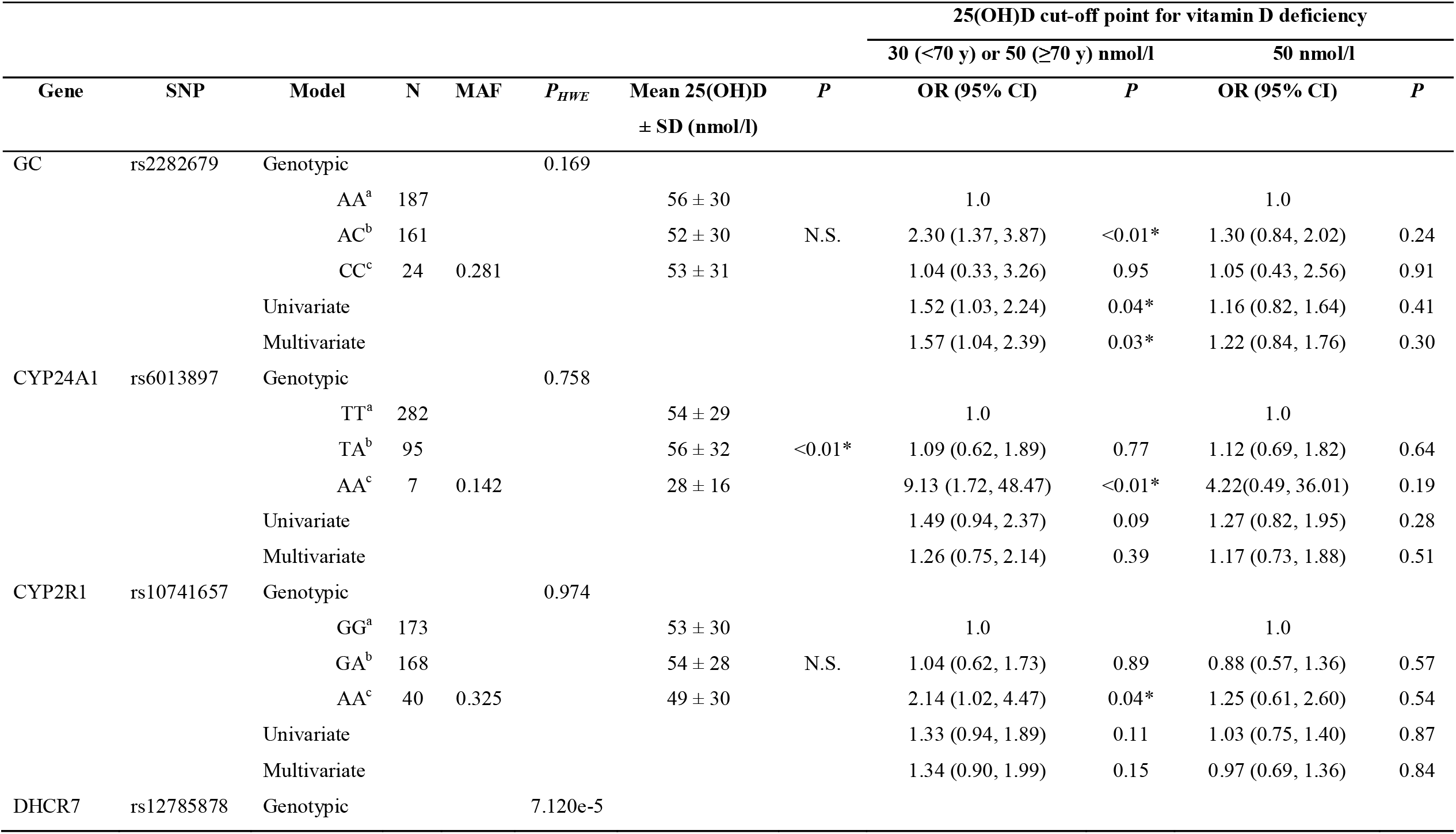
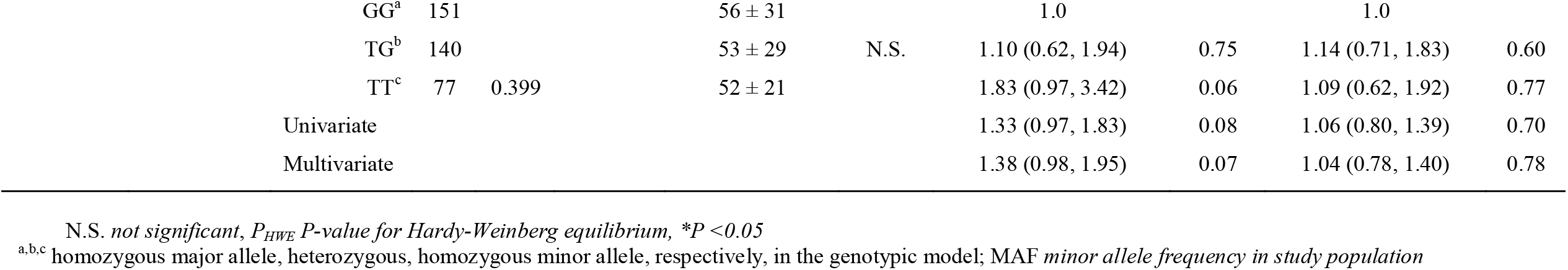
Odds ratios for the association between vitamin D genetic variants and vitamin D deficiency at two different cut-off points of 25(OH)D under a genotypic, univariate, and multivariate model (all models adjusted for age and sex; multivariate also for SNPs)

The minor allele frequency (MAF) of the C allele of rs2282679 in our study population (0.28) was in line with the frequency of 0.26 as described for East Asian populations in the 1,000 Genome Project Phase 3.

In the univariate model, the minor C allele of rs2282679 in GC gene was associated with vitamin D deficiency as defined by the criteria of the Dutch guidelines. Results of the multivariate analyses were not appreciably different from univariate analyses. However, after Bonferroni correction for four SNPs and three genotypes, no significant associations were detected anymore at the significance level of 0.004.

At the 25(OH)D cut-off point for vitamin D deficiency of 50 nmol/l, no association between any allele of the SNPs examined and vitamin D deficiency was found in either the univariate or the multivariate model (Table 2). However, a meta-analysis (N ≈ 2280) of the ORs of the C allele of rs2282679 of the GC gene in Chinese populations from Hong Kong, Dali (south-west China), and our study population, showed a significant overall OR (1.33, 95% CI 1.16, 1.52) of having vitamin D deficiency [25(OH)D <50 nmol/l] (Fig. 1).

**Fig. 1.**
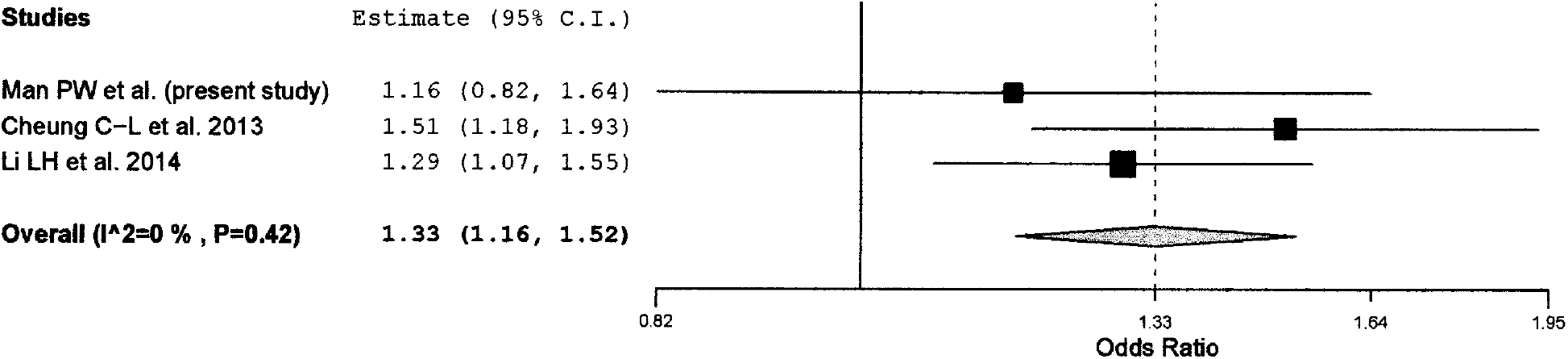
Forest plot of the C-allele of rs2282679 in GC gene associated with vitamin D deficiency [25(OH)D <50 nmol/l]

The genotypic model showed that heterozygous carriers of rs2282679 (AC) and homozygous carriers of the minor allele (AA) of rs6013897 and rs10741657 had higher risk of having vitamin D deficiency as defined by the HCN (OR = 2.30, 95% CI 1.37, 3.87; OR = 9.13, 95% CI 1.72, 48.47; OR = 2.14, 95% CI 1.02, 4.47, respectively) (Table 2). After Bonferroni correction, only heterozygous carriers of rs2282679 (AC) remained significant at the significance level of 0.004.

Serum 25(OH)D concentrations of the homozygous carriers of the major allele (TT) and the heterozygous carriers (TA) of rs6013897 were significantly higher than of the homozygous carriers of the minor allele (AA) (*P* <0.01). Of all other SNPs no difference in serum 25(OH)D concentration between homozygous carriers of the major and minor allele, and heterozygous carriers was observed (Table 2).

Results from an additionally performed linear regression analysis between the four SNPs under study and serum 25(OH)D concentration were not significant (data not shown).

## Discussion

Previous GWASs have shown that four genetic variants in or near genes involved in vitamin D transport and metabolism (rs2282679 in GC, rs6013897 near CYP24A1, rs10741657 near CYP2R1, and rs12785878 near DHCR7) can influence 25(OH)D concentration and may identify individuals with elevated risk for low concentrations of serum 25(OH)D [11-13]. The present study aimed to explore the association between these genetic variants in the vitamin D pathway and vitamin D deficiency in a sample of Chinese adults living in the Netherlands. An association was observed between the minor C allele of rs2282679 in GC and vitamin D deficiency as defined by the HCN, but this did not remain significant after Bonferroni correction.

Studies performed among Chinese Hans from Beijing and Shanghai [20], and north-eastern Han Chinese children (0-14 years) [21] showed that variants in GC and DHCR7 genes were associated with lower levels of serum 25(OH)D. The C allele of rs2282679 in GC was associated with increased risk of vitamin D deficiency [25(OH)D <50 nmol/l], as was also demonstrated in previous GWASs [11-13], in a GWAS replication study among Hong Kong Chinese women [17] and in a study performed in Dali, south-west China [18]. In the present study, the C allele of rs2282679 in GC was not significantly associated with risk of vitamin D deficiency for the 50 nmol/l cut-off, although the OR was comparable to the significant finding for the Health Council of the Netherlands recommended cut-off. Increasing power by combining studies in a meta-analysis of our findings with the aforementioned studies performed in Hong Kong and Dali, the overall OR reached the level of significance. Outside of Europe and China, an association between the C allele of rs2282679 in GC gene and vitamin D deficiency [25(OH)D <50 nmol/l] was reported in a study among 689 Mexican postmenopausal women [22]. This indicates that effects might interact with ethnicity. On the other hand, environmental influences seem less pronounced as our meta-analysis showed ORs between 1.16 and 1.51 while environmental conditions varied considerably (mainland China, Hong Kong, the Netherlands). Effect size as reported in previous GWASs varied between 1.49 [25(OH)D <50 nmol/l for vitamin D insufficiency] [13] and 1.83 [25(OH)D <25 nmol/l for vitamin D deficiency] [12], and in the aforementioned Mexican study the OR was 1.53 [25(OH)D <50 nmol/l for vitamin D deficiency], indicating some interaction between genetic background and SNP effect.

The clinical significance of SNP rs2282679 in GC gene is, however, uncertain. Although 25(OH)D will be less bound to vitamin D-binding protein due to lower concentrations of GC, it might be that the free fraction of 25(OH)D is normal in persons with this SNP. Recently, a case report study described a patient with complete vitamin D-binding protein deficiency caused by homozygous deletion of the GC gene, resulting in an apparent severe vitamin D deficiency. However, only mild bone disease (osteopenia) and fragility fractures were observed in this patient [23]. Therefore, further research in various groups is needed, examining the relationship between genetic variants in GC gene and free 25(OH)D concentration.

Regarding the other SNPs examined, results are inconsistent. One study [21] showed a positive association of CYP2R1 rs2060793 with serum 25(OH)D levels, while another study [20] could only show a trend towards an association in a Shanghai subpopulation. By contrast, two other Chinese studies [18, 24] found variants of CYP2R1 not to be significantly associated with 25(OH)D levels. Two of the aforementioned studies [20, 21] also found variants of DHCR7 associated with lower levels of serum 25(OH)D, while a study conducted in Hong Kong [17] observed no association between DHCR7 rs12785878 and serum 25(OH)D levels.

In the present study, associations between variants of CYP2R1, CYP24A1, and DHCR7 and vitamin D deficiency or serum 25(OH)D concentration were not observed. There may be several explanations for the inconsistency of study findings. First, residing in different regions in China may imply a substantial difference in genetic background [25]. In our study, we represent a diverse Chinese population [87% born in mainland China, Hong Kong or Taiwan, 7% born in the Netherlands, 6% born elsewhere; mean residence time in the Netherlands 30.5 (11.3) years] with possible different genetic and cultural backgrounds which may have influenced study results. Second, the relatively small sample size, which is a limitation of our study, may have prevented us from demonstrating small effects. Moreover, in almost 10% of our samples, DNA measurement was invalid, possibly due to technical issues with the blood collection of the participants, thereby even more reducing sample size. Third, differences in serum 25(OH)D measurement techniques may have played a role in the discrepancies in results, as different assays of serum 25(OH)D from different laboratories may not be assumed to be comparable [16].

Furthermore, in general, candidate gene studies may have several pitfalls. First, they are largely restricted by prior knowledge, and results of identified associated SNPs by candidate gene studies may not reflect the examined variant, but the effect of a nearby causal variant [26]. It could also be that genes other than those involved in transport and metabolism, but not selected due to lack of prior knowledge, had an effect on serum 25(OH)D concentrations [11]. Finally, false-positive results cannot be excluded, especially when multiple testing corrections have not been rigorously applied.

Genetic studies of vitamin D status are, nevertheless, potentially clinically relevant. It may help clinicians in counselling, screening, diagnosis, and treatment of patients with vitamin D deficiency and may be helpful in risk profiling for, e.g., osteoporosis. Therefore, in the future, it may help personalized health management as it may identify individuals or groups who would benefit most from supplementation, e.g., Chinese populations living in northern countries, or who are at risk for deficiency, which is important in view of numerous diseases associated with reduced 25(OH)D levels.

Our study has several limitations. The relatively small sample size may have prevented us from finding associations in genes other than GC. Also, possible family relationships between participants were unknown and might have influenced the calculation of *P*-values. As the study was observational, causal associations cannot be established. Moreover, our findings in the multivariate analysis should be interpreted with caution as rs12785878 near DHCR7 deviated significantly from HWE. The reason is unclear. A possible explanation is a genotyping error, although no problems during DNA isolation or genotyping could be identified. Furthermore, our meta-analysis was on the basis of only three observational studies, which carries the risk of bias. However, all studies adjusted for age, and two out of the three studies also adjusted for sex; the third study only included women. Moreover, the odds ratios were of very comparable magnitude. Finally, our findings are not generalizable to other Chinese populations as genetic make-up may differ among Chinese depending on population structure and geographic origin of the participants [25]. Strength of our study was that no influence of season was present, as all measurements were performed in March 2014. In conclusion, this study found the C allele of rs2282679 in GC gene associated with vitamin D deficiency. Associations between the other SNPs and vitamin D deficiency or between SNPs and serum 25(OH)D concentration were not observed.

## Data Availability

The data that support the findings of this study are available from the corresponding author upon reasonable request.

## Acknowledgments

The authors thank all responders for their participation and all volunteers for their efforts. The authors gratefully acknowledge financial support from ‘Stichting Artrose Zorg’ and ‘Fonds voor het Hart’. Also, the authors thank Dr. Li-Hua Li (Dali University School of Clinical Medicine, China) for sharing their data, Professor André Uitterlinden (Department of Internal Medicine, Erasmus MC, University Medical Center, Rotterdam, the Netherlands) for proofreading the manuscript and Ramazan Buyukcelik and Pascal Arp (Department of Internal Medicine, Erasmus MC, University Medical Center, Rotterdam, the Netherlands) for technical assistance.

## Author contributions

P.W.M., study design, data collection, data analysis, manuscript preparation; S.B., data analysis, manuscript preparation; E.J.F.H., data analysis, manuscript preparation; W.L., data collection, manuscript preparation; M.E.N., study design, manuscript preparation; P.L., study design, data analysis, manuscript preparation; B.J.C.M., study design, data analysis, manuscript preparation. All authors revised the manuscript for important intellectual content.

